# External validation of a decision rule for bacteremia vs contaminants in pediatric blood cultures

**DOI:** 10.64898/2026.07.17.26358300

**Authors:** Madeleine D’amours-Gravel, Alessia Charvet, Carla Ibañez Miguel, Nine Rouxel, Camille Fontaine, Julie Besson, Léna Jiguet, Liam Karara, Luna Pozzi, Claudia Teixeira, Carla Henoud-Bertaina, Céline Alves, Abdessalam Cherkaoui, Delphine S. Courvoisier, Johan N. Siebert

## Abstract

**BACKGROUND:** Half of positive blood cultures in pediatric emergency departments (PEDs) represent contaminants, driving unnecessary hospitalization, antibiotic exposure, and repeat visits. A clinical decision rule derived at CHU Sainte-Justine showed 99% sensitivity and 60% specificity for distinguishing bacteremia from contaminants but had not been externally validated. We sought to validate this rule in an independent pediatric cohort.

**METHODS:** This retrospective diagnostic study spanned from January 2015 to May 2025 at a tertiary PED in Switzerland, using positive blood cultures from patients younger than 16 years. The four predictors (Gram-negative organisms or Gram-positive cocci in pairs or chains; time to positivity <17 hours; indwelling device; suspected osteoarticular infection) classified each case as low, moderate, or high risk. The primary outcome was bacteremia, adjudicated by two independent reviewers, based on organism identity and infectious disease specialist’s assessment. Diagnostic accuracy was assessed with 95% CIs.

**RESULTS:** Of 130 children enrolled (median age 3.8 years [IQR 0.9-9.9]; 61.5% male), 78 (60.0%) had true bacteremia. The rule yielded a sensitivity of 97.4% (95% CI, 91.0-99.7), specificity of 69.2% (95% CI, 54.9-81.3), positive predictive value of 82.6% (95% CI, 73.3-89.7), and negative predictive value of 94.7% (95% CI, 82.3-99.4). Both false-negatives were immunocompetent children with methicillin-susceptible *Staphylococcus aureus* bacteremia without indwelling devices. Among contaminants, 71% received antibiotics under usual care versus 31% classified as moderate or high risk by the rule.

**CONCLUSIONS:** This first external validation supports the Sainte-Justine rule in a distinct pediatric population, preserving sensitivity with higher specificity. Multicenter validation is warranted before adoption.

## 1. Introduction

### 1.1 Background

Fever accounts for 10 to 20% of pediatric emergency department (PED) visits and is the most common chief complaint among infants and children [1, 2]. Bacteremia is the underlying cause in 1.5% to 2.3% of these patients [3]. Blood cultures are routinely obtained in febrile children, where guidelines strongly support their use [4, 5], and in those with suspected invasive bacterial infection [6, 7]. Although timely recognition of true bacteremia is critical given the risk of progression to meningitis, septic shock, metastatic infections, or death [8–10], 40% to 60% of positive blood cultures represent contaminants [11, 12]. This figure reflects a low underlying prevalence of true bacteremia rather than high absolute contamination rates, which remain in the 1.5 to 6.5% range across all PED blood cultures [13, 14].

### 1.2 Importance

False-positive results carry measurable clinical and economic consequences [15]. Children with contaminated cultures often return to the PED, undergo additional venipunctures, are admitted unnecessarily, and receive empirical antibiotics, with attendant risks of adverse events, line-related complications, and resistance selection [4, 12, 13, 16, 17]. A multicenter survey confirmed substantial management heterogeneity. In identical scenarios involving a positive blood culture, 49% of respondents would have discharged the patient whereas 35% would have admitted for intravenous antibiotics [18]. This variability persists despite American Academy of Pediatrics guidance [4] and the PECARN febrile-infant rule [19], neither of which addresses the management of a positive blood culture once obtained.

Real-time molecular diagnostics, including multiplex polymerase chain reaction (PCR), syndromic panels, and metagenomic next-generation sequencing are increasingly used alongside conventional blood cultures and can accelerate pathogen identification and antimicrobial optimization. Their contribution to the specific task of separating true bacteremia from contamination is, however, limited. Their targeted panels miss organisms not included in the assay, with recognized coverage gaps in pediatric populations [20]. Their high analytical sensitivity detects nucleic acid from both viable and non-viable organisms, complicating discrimination of true pathogens [21]. And their results require careful correlation with the clinical presentation and other microbiological findings [22]. High cost of instruments, reagents, and maintenance further limits widespread implementation [23].

Several predictors discriminate true bacteremia from contaminants, including Gram-stain morphology, time to positivity (TTP), inflammatory markers, indwelling device, and suspected osteoarticular infection [11, 24–27]. TTP appears particularly informative, with shorter values for true bacteremia and 90% of case become positive within 24 hours [25]. Procalcitonin and C-reactive protein, although useful for invasive bacterial infections in young febrile infants [28, 29], are of limited value for this specific distinction. Combining such predictors into a standardized decision tool is therefore the logical next step. The Hospital for Sick Children algorithm, externally validated in Montreal, achieved 100% sensitivity but only 11% specificity [11], an imbalance that limits clinical utility. More recently, Gravel et al. derived and internally validated a four-predictor clinical decision rule (CDR) in children with positive blood cultures, yielding 99% sensitivity and 60% specificity [24].

### 1.3 Goals of this investigation

Because clinical prediction models require external validation in independent settings before safe and generalizable adoption [30, 31], and none has been published for this rule, our objective was to perform the first external validation of the Sainte-Justine four-predictor CDR in an independent pediatric cohort.

## 2. Methods

### 2.1 Study design and setting

We conducted a retrospective, single-center cohort study from January 1, 2015 to May 31, 2025 in the tertiary PED of the Geneva University Hospitals (HUG), Switzerland, managing approximately 36,000 visits per year. The bacteriology laboratory processes blood cultures using continuous-monitoring automated incubation BD BACTEC™FX systems that flag positivity 24 hours per day. In 2024, 78,768 blood culture bottles were submitted (positivity rate 6.2%) [32], with delayed loading over 24 hours in 0.9%. Laboratory operations run from 7:30 A.M. daily, closing at 7:30 P.M. on weekdays, 5:00 P.M. on Saturdays, and 1:00 P.M. on Sundays. Ethical approval was obtained from the Geneva Research Ethics Commission (CCER, approval number: 2025–01069) and written informed consent was obtained for all research individuals. The study was in accordance with the Helsinki Declaration [33] and followed applicable reporting guidelines [30, 34, 35].

### 2.2 Participants

#### Inclusion criteria

Children younger than 16 years at the PED visit, with at least one positive blood culture drawn during that visit, and written informed consent for secondary use of medical data from a legal representative or the participant.

#### Exclusion criteria

Age 16 years or older at the index visit; negative or inconclusive blood cultures; absence of a recorded TTP value or other incomplete records for variables required to apply the CDR; and ambiguous diagnostic information unresolved by independent adjudication.

Eligible cases were identified through the institutional microbiology database and cross-referenced against the electronic health record (EHR). To minimize selection bias, they were randomly ordered before screening and enrollment continued until the prespecified sample size was achieved. Because recruitment was capped at the target sample size, positive cultures not screened by that point were not assessed. Enrolled patients therefore represented a random subset of all eligible cases rather than a clinically selected sample.

### 2.3 Outcomes

The primary outcome was true bacteremia, defined with the same two-step adjudication approach used in the derivation study [24], enabling direct comparability between cohorts. The reference-standard was established independently and prior to the CDR result. For each positive culture, two independent reviewers (MDG, JNS) classified the result as true bacteremia or contaminant on the basis of 1) the identity of the organism, categorized using the Centers for Disease Control and Prevention list of common commensals [36] and the reference categorization of probable contaminants [37], and 2) the pediatric infectious disease specialist’s report documented in the EHR. Disagreements were resolved by consensus, with a third reviewer available for arbitration. The four CDR predictors were extracted and the risk classification was computed only afterward. C-reactive protein and procalcitonin were recorded as descriptive variables only and contributed neither to the reference-standard adjudication nor to the CDR classification.

### 2.4 Predictors

The four CDR predictors were derived by the original authors from a literature review and expert consensus, then selected through Classification and Regression Tree modeling optimized for sensitivity. We applied them in the sequential structure (Fig 1) proposed by Gravel et al. [24]:

- Gram-negative organisms OR Gram-positive cocci arranged in pairs or chains on initial Gram stain (high-risk gate).
- TTP below 17 hours from placement of the bottle into the BD BACTEC™FX system to detection of microbial growth.
- Indwelling devices (S1 Table).
- Clinical suspicion of osteoarticular infection (septic arthritis or osteomyelitis at the index visit).

**Fig 1.**
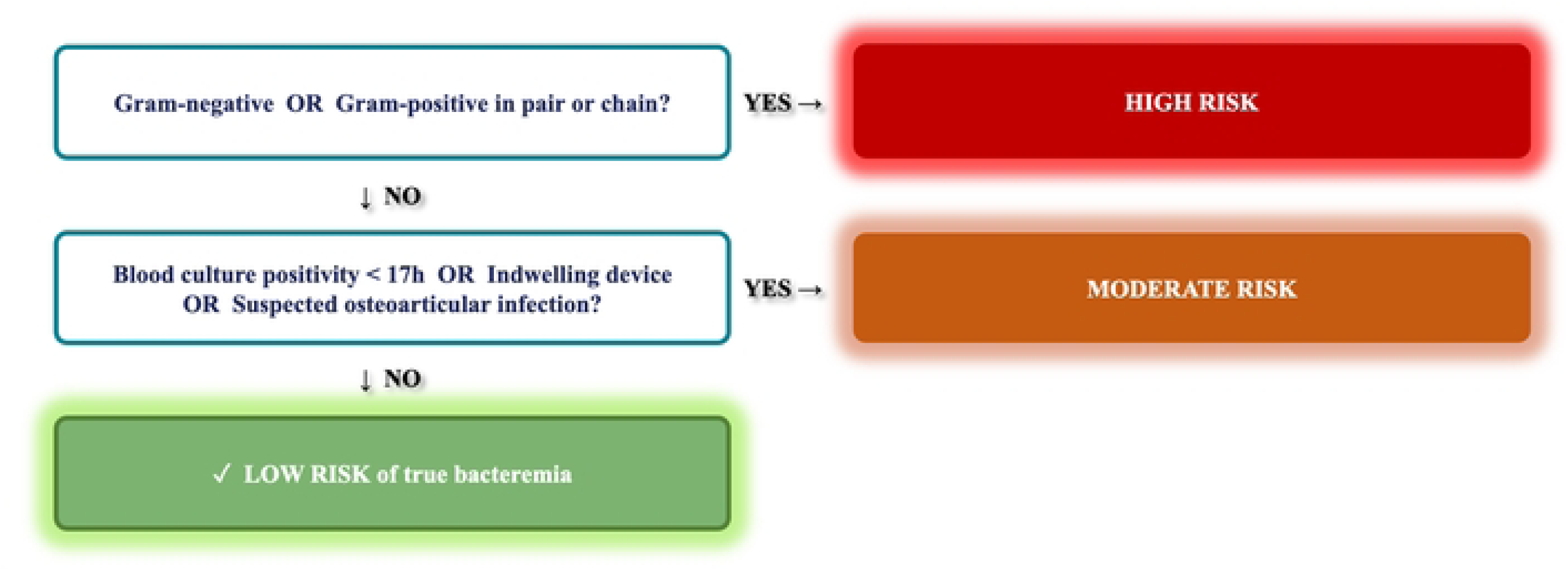
The Sainte-Justine Clinical Decision Rule for Pediatric Positive Blood Cultures. Sequential application of the four predictors classifies patients into three risk strata. Patients meeting the Gram-stain criterion (Gram-negative bacteria or Gram-positive organisms in pairs or chains) are classified as high risk. Among the remainder, those with at least one of the three secondary criteria (indwelling device, suspicion of osteoarticular infection, or TTP < 17 hours) are classified as moderate risk. Patients meeting none of the four criteria are classified as low risk of true bacteremia. Adapted with permission from Gravel et al [24].

### 2.5 Data Extraction

Each chart was independently reviewed by two investigators (MDG, JNS) using a structured case report form derived from the original Sainte-Justine instrument. Extracted variables included demographics, comorbidities, presenting features, laboratory and microbiological variables, and management data (S2 Table).

### 2.6 Sample Size

Based on a true bacteremia prevalence of approximately 50% in positive pediatric blood cultures in the derivation study [24], we estimated that 130 evaluable cases would provide at least 64 true bacteremia cases. This is consistent with established recommendations for external validation of clinical prediction models [31, 38] and allows estimation of sensitivity with a lower 95% confidence interval bound of approximately 0.94, given an expected point estimate of 0.99.

### 2.7 Statistical Analysis

Continuous variables are reported as median (interquartile range [IQR]) and were compared using the Mann-Whitney U test. Differences in medians were estimated using the Hodges-Lehmann estimator with 95% confidence intervals (CIs). Categorical variables are reported as number (percentage) and were compared using the Fisher exact test. Differences in proportions are reported with 95% CIs calculated using the Newcombe method with continuity correction. For Gram-stain morphology, a single global comparison of the distribution of the six morphotypes between groups was performed using the Fisher-Freeman-Halton exact test. The four CDR predictors were applied to each case and the resulting risk classification was compared with the reference standard. Diagnostic accuracy was assessed by sensitivity, specificity, positive predictive value (PPV), and negative predictive value (NPV), with Clopper-Pearson exact 95% CIs. The proportion of true bacteremia within each TTP interval was reported with Wilson score 95% CIs. To assess abstraction reliability, a random 10% of charts was independently reviewed by a third investigator (CIM) blinded to the initial extraction. Interrater agreement was assessed using Cohen’s κ for the two variables requiring interpretive judgment: the reference-standard outcome (true bacteremia versus contaminant) and suspected osteoarticular infection. The remaining predictors (Gram-stain result, TTP, indwelling device) were extracted directly from laboratory reports or medical records and were not subject to formal reliability assessment. As a secondary analysis, we estimated the antibiotic exposure that would have been avoided by applying the CDR to discharge low-risk cases without antibiotics, using observed prescribing patterns as the comparator. As a sensitivity analysis to assess the robustness of the rule to the choice of TTP threshold, we recalculated diagnostic accuracy while varying the TTP cutoff continuously from 12 to 24 hours, holding the three other predictors constant. Sensitivity, specificity, and predictive values with Clopper-Pearson exact 95% CIs were derived at each threshold and are presented graphically across the full range and tabulated at hourly increments. All tests were two-sided, and a P value less than 0.05 was considered statistically significant. No correction for multiple testing was applied given the study’s pre-specified primary endpoint. Analyses were performed using R version 4.6.0 (R Foundation for Statistical Computing).

## 3. Results

### 3.1 Study Population

From January 1, 2015 to May 31, 2025, 324 positive blood cultures were identified, and 130 evaluable cases were enrolled (Fig 2). Their demographic, clinical, and microbiological characteristics stratified by reference-standard outcome (true bacteremia versus contaminant) are presented in Table 1. Immunocompromised status did not differ significantly (15.4% vs 26.9%, *P* = 0.137), whereas indwelling devices and suspected osteoarticular infections were strongly associated with true bacteremia (both *P* < 0.001). C-reactive protein and procalcitonin were markedly higher in true bacteremia (both *P* < 0.001). Interrater agreement was κ = 1.00 for both the reference-standard outcome and the suspected osteoarticular infection predictor across the 13 randomly sampled duplicate charts. The corresponding characteristics of the original validation cohort [24] are presented in S3 Table.

**Fig 2.**
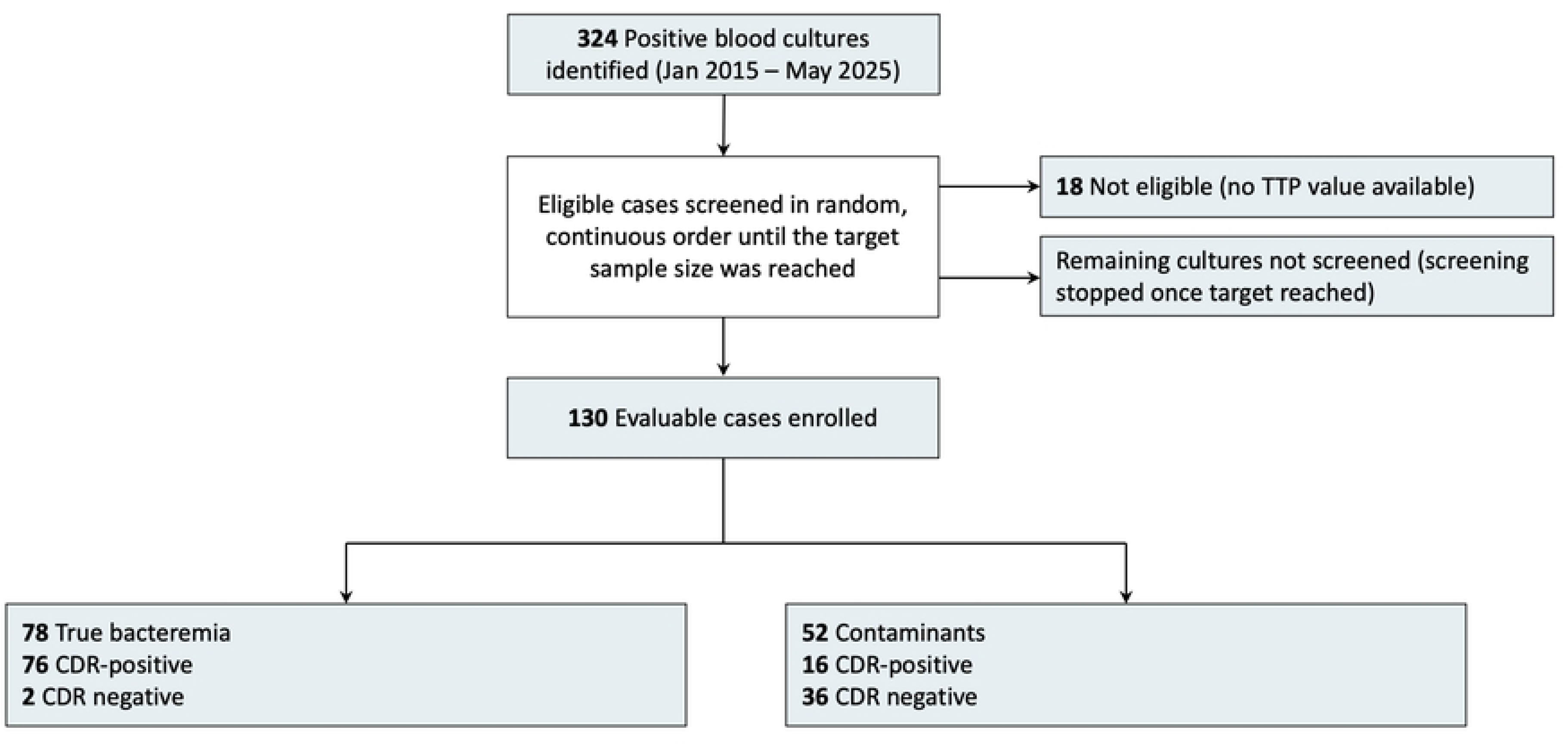
Flow of participants through the validation cohort.

**Table 1.**
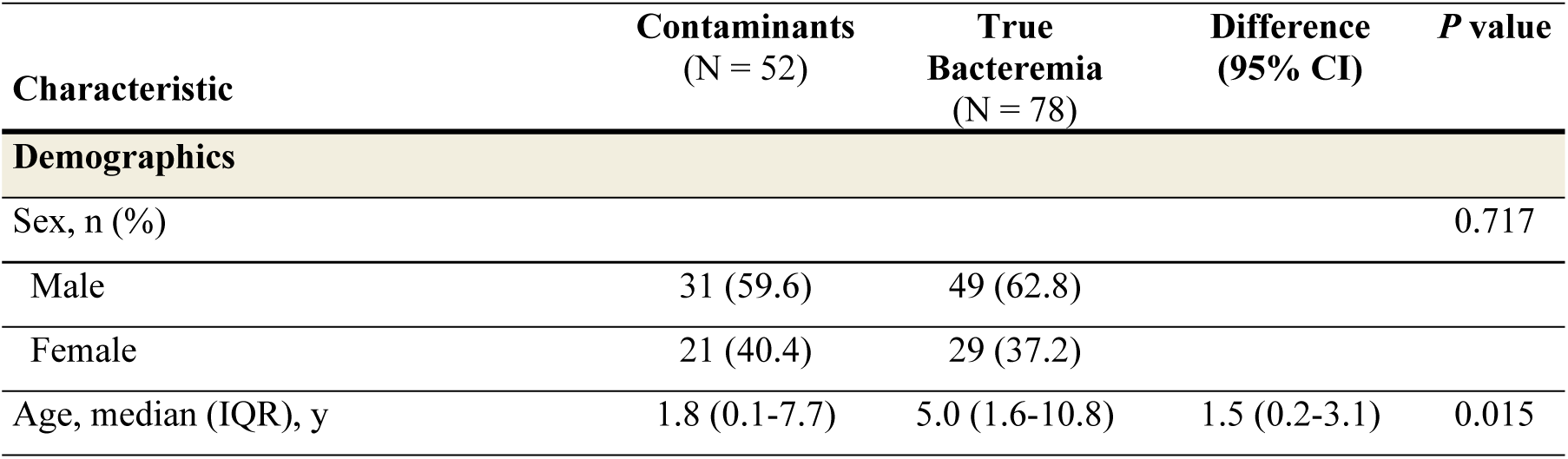
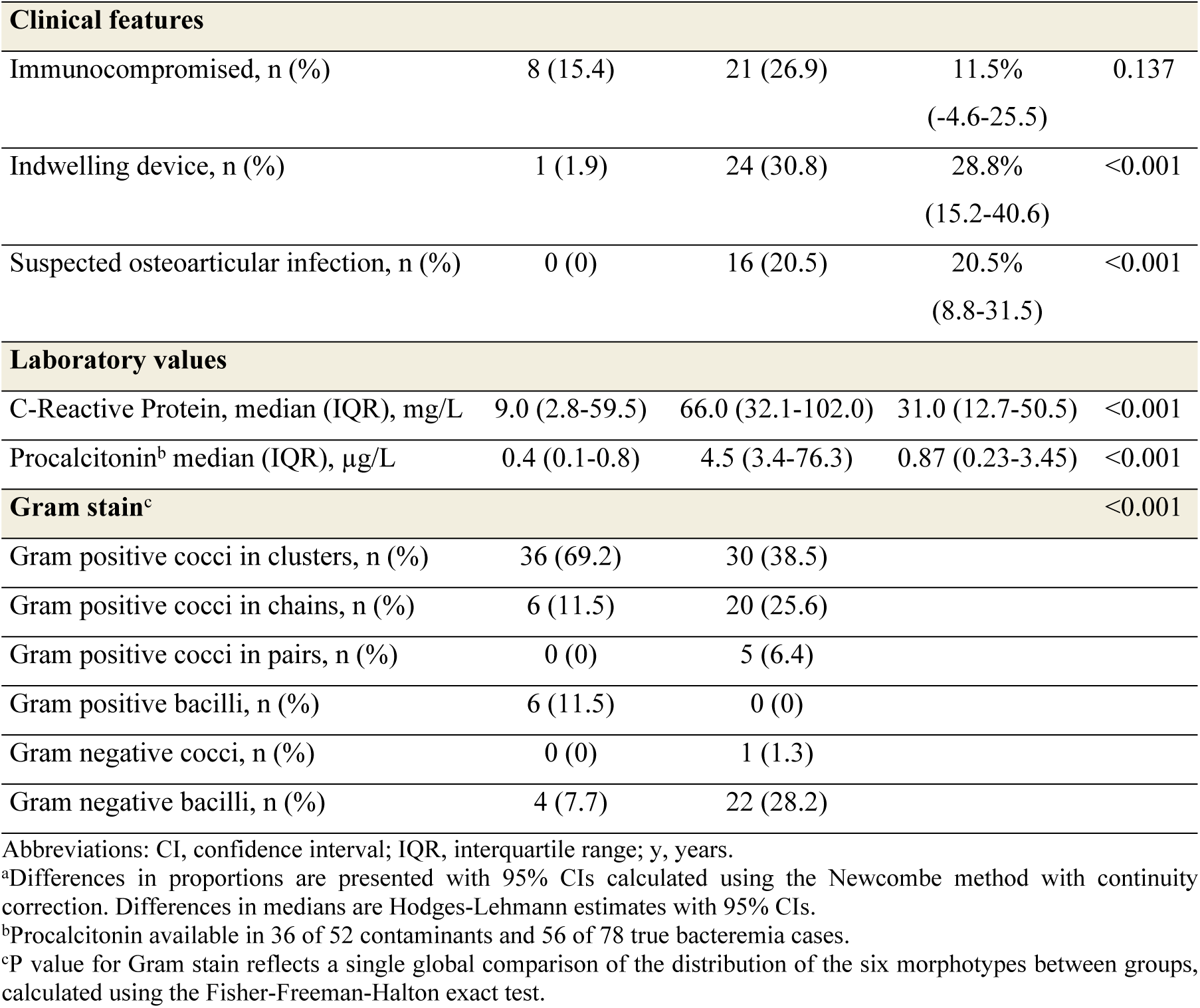
Baseline and Clinical Characteristics of Study Patients, by Reference-Standard Classification.

### 3.2 Main results

The median TTP was 11.8 hours (IQR, 8.3-16.9) for true bacteremia and 22.8 hours (IQR 17.8-33.5) for contaminants, a difference of -11.0 hours (95% CI, -12.9 to -7.8). The proportion of true bacteremia declined as TTP increased, from 24 of 27 cases (88.9%; 95% CI, 71.9%-96.1%) within 10 hours to 3 of 14 (21.4%; 95% CI, 7.6%-47.6%) by 22 to 24 hours (Fig 3). Overall, 72 of 78 true bacteremias (92.3%; 95% CI, 84.2%-96.4%) were detected within 24 hours of sampling.

**Fig 3.**
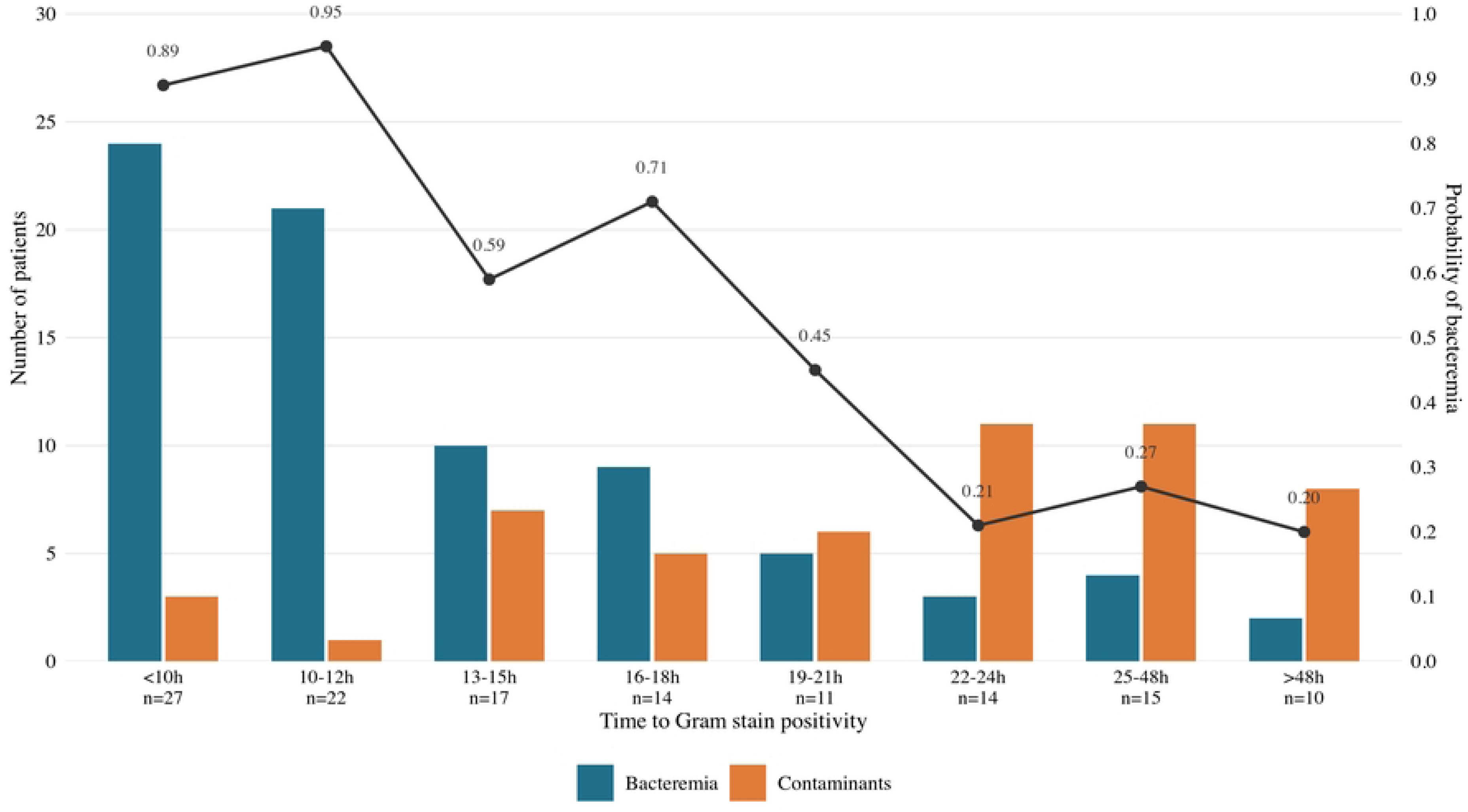
Distribution of bacteremia vs. contaminants according to TTP (N = 130)

### 3.3 Performance of the Sainte-Justine CDR

Of the 78 children with true bacteremia, the CDR classified 48 (61.5%) as high risk and 28 (35.9%) as moderate risk. Two were classified as low risk (false negatives). Of the 52 children with contaminants, 36 (69.2%) were correctly classified as low risk, while 10 (19.2%) were classified as high risk and 6 (11.5%) as moderate risk (false positives). Overall diagnostic performance is provided in Fig 4.

**Fig 4.**
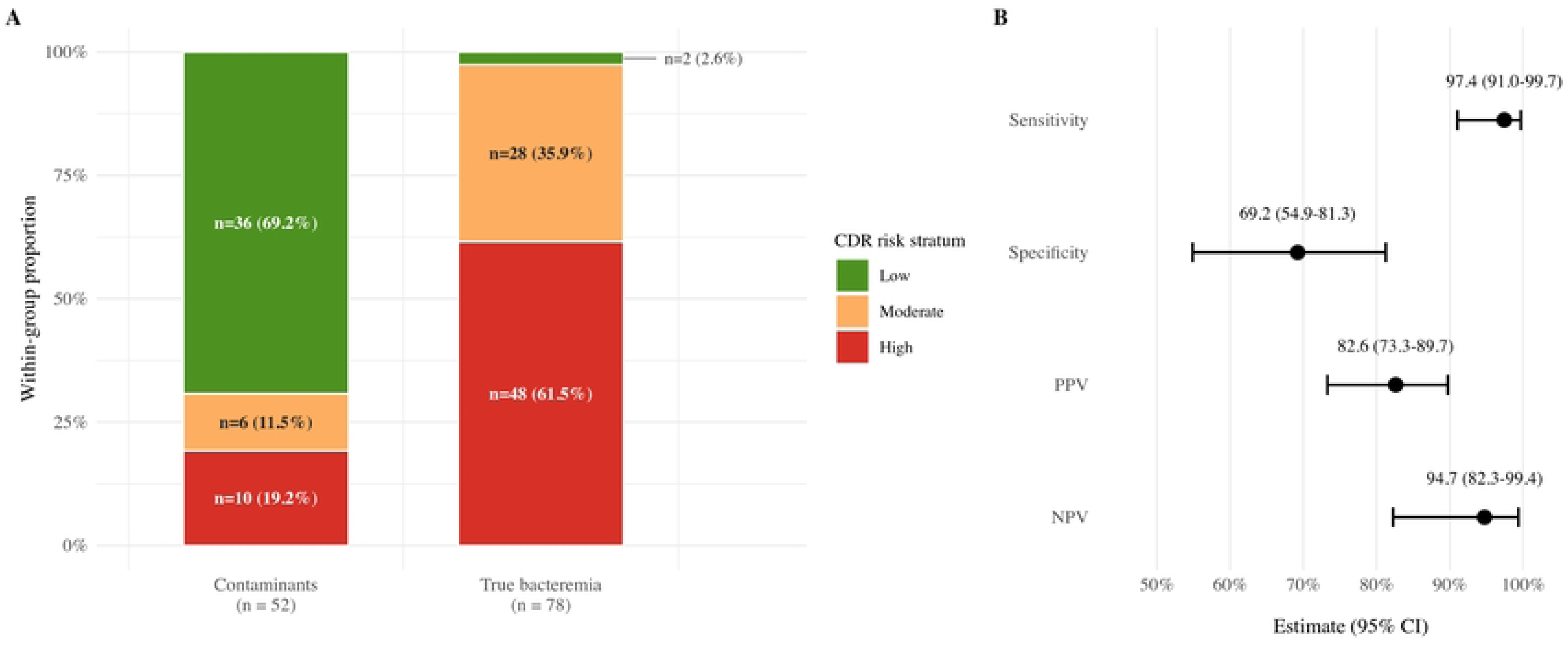
Risk stratification and diagnostic performance of the clinical decision rule for distinguishing true bacteremia from contaminants. (A) Distribution of cases across the three CDR risk strata. (low, moderate, high), shown separately for contaminants (N = 52) and true bacteremia (N = 78). Bars represent the within-group proportion, with absolute counts and percentages labeled. The gradient illustrates the rule’s discrimination, with most contaminants classified as low risk and most true bacteremia as high risk. (B) Diagnostic accuracy of the rule with point estimates and 95% confidence intervals for sensitivity, specificity, positive predictive value, and negative predictive value. Confidence interval calculated by the Clopper-Pearson exact method. CDR, clinical decision rule; CI, confidence interval; NPV, negative predictive value; PPV, positive predictive value.

### 3.4 Threshold sensitivity analysis

Across the full range from 12 to 24 hours, sensitivity remained consistently high (94.9% to 98.7%), as did the negative predictive value (91.1% to 96.8%). As expected, raising the threshold increased sensitivity at the cost of specificity, which declined from 78.8% (95% CI, 65.3-88.9) at 14 hours to 57.7% (95% CI, 43.2-71.3) at 20 hours (Fig 5 and S4 Table). At the derivation threshold of 17 hours, sensitivity was 97.4% (95% CI, 91.0-99.7) and specificity 69.2% (95% CI, 54.9-81.3).

**Fig 5.**
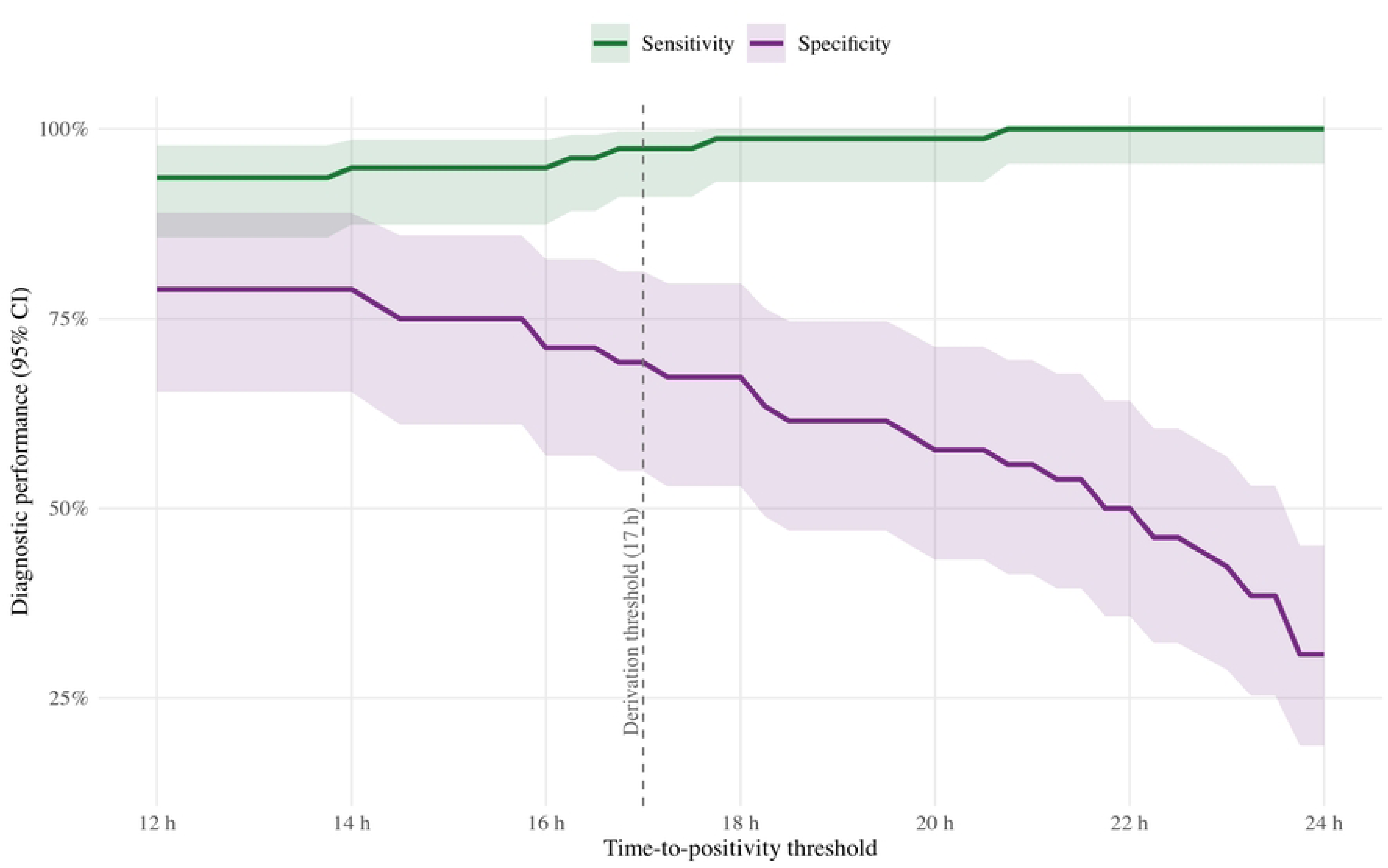
Sensitivity (green) and specificity (purple) of the clinical decision rule across time-to-positivity thresholds (12–24 hours). Shaded bands are 95% confidence intervals. The dashed line marks the 17-hour derivation threshold.

### 3.5 False negative cases

Both children misclassified as low risk had confirmed true bacteremia caused by methicillin-susceptible *Staphylococcus aureus.* One isolate was Panton-Valentine leukocidin-positive. Neither patient was immunocompromised or carried an indwelling device. In both, the Gram stain did not fulfil the high-risk morphology criterion, and TTP exceeded the 17-hour threshold (17.7 and 20.6 hours, respectively). These cases therefore represent borderline presentations in which classification was driven solely by a TTP value above the cut-off, with no other high-risk feature.

### 3.6 Potential impact on antibiotic exposure

In the contaminant group, 37 of 52 children (71.2%) received antibiotics under usual practice. Of these 37, 26 (70.3%) were classified as low risk by the CDR and represent potentially avoidable antibiotic exposure. Applying the rule would thus have reduced antibiotic exposure among contaminants from 71.2% to 21.2%, an absolute reduction of 50 percentage points, corresponding to one unnecessary antibiotic course potentially averted for every two contaminants to whom the rule was applied. The two false-negative cases received antibiotics regardless of CDR classification, reflecting the role of clinical judgement at the bedside.

## 4. Discussion

### 4.1 Main findings and interpretation

This first external validation of the Sainte-Justine CDR in a Swiss tertiary pediatric emergency cohort confirms strong diagnostic performance. The sensitivity of 97.4% is marginally lower than the 99% at derivation [24], consistent with the accuracy loss commonly seen when prediction models are transported to new settings [31, 38]. The specificity of 69.2% was higher than the 60% reported by Gravel et al. [24] and the 11% of the Hospital for Sick Children algorithm evaluated in the same Montreal cohort [11]. The management heterogeneity reported in a recent Canadian survey [18] reflects a genuine unmet need for structured decision support.

### 4.2 Interpretation of false negatives

The two missed cases warrant scrutiny, as sensitivity is the principal safety property of a rule that defers antibiotics. Both occurred in immunocompetent children without an indwelling device, and in both the Gram stain did not fulfil the high-risk morphology criterion, with. TTP just above the threshold. These values are consistent with published pediatric distributions for *Staphylococcus aureus* bacteremia, which extend well beyond 17 hours and are substantially longer than for pathogens such as *Escherichia coli* or group B *Streptococcus* [39, 40]. Both cases are therefore microbiologically plausible as true bacteremia, although no firm conclusions can be drawn from two observations. Critically, neither presented any other high-risk feature that could have served as a safety signal at culture reporting. This argues for pairing rule-based antibiotic deferral with a structured 24-hour clinical and microbiological safety net, rather than pursuing threshold refinement alone.

The TTP predictor depends on pre-analytical conditions. Delayed loading of bottles held at room temperature substantially shortens TTP [32]. TTP is therefore modulated by transport time, holding temperature, and laboratory hours, rather than being solely intrinsic to the organism. At our institution, short transport times and infrequent delayed loading made such shortening unlikely [32], but the 17-hour threshold may not transfer unchanged across laboratories with different pre-analytical workflows, supporting local calibration in any prospective implementation. The practical impact of this dependence, however, hinges on how much the rule’s accuracy actually shifts when the threshold is varied, which we examined directly.

### 4.3 Clinical decision rule robustness

Our sensitivity analysis showed that performance was stable across TTP thresholds from 12 to 24 hours, with sensitivity remaining above 94% throughout. This robustness is clinically reassuring, because the optimal TTP cutoff reported in the literature varies appreciably with the population, the blood culture system, and the era of testing. Recent studies have proposed thresholds of approximately 16 to 20 hours, reflecting the shorter detection times of modern automated systems [41, 42]. The derivation threshold of 17 hours falls within this range, and the absence of any abrupt change around it indicates that the rule does not depend on precise calibration to a single institution’s culture system. This is consistent with the underlying biology. Most bacteremias are detected early, with roughly 90% positive by 24 hours [39], so the rule retains high sensitivity even when the threshold is lowered.

### 4.4 Molecular diagnostics and the persistent need for contaminant discrimination

The growing availability of rapid molecular diagnostics might appear to reduce the need for a clinical rule. These assays, however, address a different question. These panels accelerate organism identification and antimicrobial optimization once a culture is positive, but do not determine whether the detected organism represents true infection or contamination [43]. Their higher analytical sensitivity may in fact even increase detection of contaminant DNA. In one pediatric study, PCR yielded contaminant results in 22% of samples versus 6% for culture [44]. A recent systematic review and meta-analysis found that rapid molecular assays still lack the sensitivity to replace blood culture as a standalone diagnostic [45]. Their diagnostic performance improves when interpreted against a composite clinical diagnosis [46, 47]. The task of discriminating true bacteremia from contaminants therefore remains fundamentally clinical, and a structured rule complements rather than competes with molecular methods.

### 4.5 Comparison with previous works

Compared with the Hospital for Sick Children algorithm [11], the Sainte-Justine CDR achieves a more favorable balance between sensitivity and specificity, largely because the earlier algorithm classified most contaminants as bacteremia. The higher specificity reflects in part the inclusion of TTP as a predictor, consistent with the reported association between rapid positivity and true bacteremia [25]. These comparisons warrant caution, as sensitivity and specificity remain sensitive to differences in case mix and severity across cohorts, and the TTP threshold is itself subject to pre-analytical variability [48].

Our findings also align with evidence on the limitations of biomarkers as standalone discriminators. Neither C-reactive protein nor procalcitonin, nor composite tools such as the Lab-score were designed for the binary task of distinguishing true bacteremia from contamination. Both perform moderately well for severe bacterial infection, but were validated in broader febrile populations rather than in children with a preliminary positive blood culture [28, 29, 49]. Structured rules combining microbiological, clinical and host predictors are therefore more likely than biomarkers alone to support management changes in this scenario.

### 4.6 Clinical implications

If reproduced prospectively, the rule could reduce unnecessary antibiotic exposure while preserving the safety net for true bacteremia. In our cohort, 71% of children with contaminant cultures received antibiotics under usual practice. Half of all contaminants (26 of 52) were both treated and classified as low risk by the CDR, representing antibiotic courses that could potentially have been avoided. Antibiotic stewardship in pediatric sepsis workup remains a recognized priority, and structured discrimination of positive blood cultures offers a concrete entry point for bedside stewardship.

### 4.7 Limitations

This study has several limitations. First, the reference standard was not fully independent from the predictors included in the CDR, particularly Gram-stain morphology and clinical suspicion of osteoarticular infection, creating a risk of incorporation bias that may have overstated observed diagnostic performance. But no perfect reference standard exists for contamination. These risks were mitigated by blinded sequential adjudication, two independent reviewers, and a structured case report form. Second, the specialist’s assessment incorporated the Gram-stain result and organism identity, which also inform the CDR’s high-risk gate, raising potential incorporation bias for that predictor. However, TTP, the rule’s most discriminant predictor, was not available to clinicians at the bedside and could not have influenced the reference standard. Third, the sample size of 130 evaluable cases yields acceptable but relatively wide confidence intervals, particularly for specificity, although the sensitivity analysis confirmed that diagnostic performance was robust across TTP thresholds from 12 to 24 hours. Fourth, single-center recruitment limits generalizability. Fifth, the antibiotic exposure estimate reflects observed prescribing patterns rather than prospective behavior, and random case selection minimizes selection bias. Finally, in our cohort, 61 of 130 cultures (46.9%) were collected outside laboratory operating hours and remained at room temperature before loading into the continuous-monitoring automated incubation systems, potentially resulting in shorter measured TTP values. As this effect would shift cases toward the high-risk side of the 17-hour threshold, it would be expected to maintain rather than compromise the rule’s sensitivity [32].

## 5. Conclusion and perspectives

This first external validation of the Sainte-Justine CDR supports its diagnostic performance. The rule offers a more favorable balance between sensitivity and specificity than the Hospital for Sick Children algorithm and could reduce unnecessary antibiotic exposure in pediatric emergency settings. Prospective multicenter validation within pediatric emergency research networks is the next step. Impact studies should quantify changes in antibiotic prescribing, length of stay, return visits, and missed bacteremia, and determine whether the TTP threshold requires local calibration given institutional differences in pre-analytical workflows, and whether a structured 24-hour follow-up pathway is needed when antibiotics are deferred.

## Data Availability

The data consist of individual patient records extracted from the electronic health record of Geneva University Hospitals, Switzerland. Individual-level data cannot be shared publicly because they are sensitive patient data, and public release is precluded by the terms of the ethics approval and patient consent and by the risk of reidentification in a single-center pediatric cohort. The datasets generated during and/or analysed during the current study are available in deidentified form from the corresponding author on reasonable request.

## 6 Acknowledgements

We thank the PIEUVRE (Program for the Integration of Early-career University student Volunteers in RESearch) team at Geneva University Hospitals for their support throughout this work, and the microbiology laboratory of HUG for facilitating data extraction.

## 8. Supporting information

S1 Table. Indwelling devices

S2 Table. Data Extraction

S3 Table. Baseline and Clinical Characteristics of the Sainte-Justine Derivation Cohort

S4 Table. Diagnostic Accuracy of the Clinical Decision Rule Across Time-to-Positivity Thresholds From 12 to 24 Hours

